# SARS-CoV-2 Omicron VOC Transmission in Danish Households

**DOI:** 10.1101/2021.12.27.21268278

**Authors:** Frederik Plesner Lyngse, Laust Hvas Mortensen, Matthew J. Denwood, Lasse Engbo Christiansen, Camilla Holten Møller, Robert Leo Skov, Katja Spiess, Anders Fomsgaard, Maria Magdalena Lassaunière, Morten Rasmussen, Marc Stegger, Claus Nielsen, Raphael Niklaus Sieber, Arieh Sierra Cohen, Frederik Trier Møller, Maria Overvad, Kåre Mølbak, Tyra Grove Krause, Carsten Thure Kirkeby

## Abstract

The Omicron variant of concern (VOC) is a rapidly spreading variant of SARS-CoV-2 that is likely to overtake the previously dominant Delta VOC in many countries by the end of 2021.

We estimated the transmission dynamics following the spread of Omicron VOC within Danish households during December 2021. We used data from Danish registers to estimate the household secondary attack rate (SAR).

Among 11,937 households (2,225 with the Omicron VOC), we identified 6,397 secondary infections during a 1-7 day follow-up period. The SAR was 31% and 21% in households with the Omicron and Delta VOC, respectively. We found an increased transmission for unvaccinated individuals, and a reduced transmission for booster-vaccinated individuals, compared to fully vaccinated individuals. Comparing households infected with the Omicron to Delta VOC, we found an 1.17 (95%-CI: 0.99-1.38) times higher SAR for unvaccinated, 2.61 times (95%-CI: 2.34-2.90) higher for fully vaccinated and 3.66 (95%-CI: 2.65-5.05) times higher for booster-vaccinated individuals, demonstrating strong evidence of immune evasiveness of the Omicron VOC.

Our findings confirm that the rapid spread of the Omicron VOC primarily can be ascribed to the immune evasiveness rather than an inherent increase in the basic transmissibility.

## 2 Introduction

The SARS-CoV-2 variant B.1.1.529, which is referred to as the Omicron variant of concern (VOC), has overtaken the Delta VOC in South Africa and has spread rapidly to at least 28 countries countries in Europe (7), Asia, the Middle East and South America (9; 17). The Omicron VOC has been reported to be three to six times as infectious as previous variants (4), with a short doubling time (11), including early estimates from countries with a high vaccination coverage indicating doubling times of 1.8 days (UK), 1.6 days (Denmark), 2.4 days (Scotland) and 2.0 days (United States) (26). Transmission of the Omicron VOC has been high among individuals being fully vaccinated against SARS-CoV-2 infection as well as among individuals with a history of COVID-19 infection (19).

A current concern worldwide is that the Omicron VOC is able to evade immunity induced by the currently used vaccines, and a preliminary meta-analysis of neutralization studies indicated that the vaccine effectiveness is reduced to around 40% against symptoms and to 80% against severe disease, but that the effect for booster vaccinations is at 86% and 98%, respectively (12). These results are supported by laboratory studies establishing a markedly reduced elimination of the Omicron VOC by neutralizing antibodies, indicating that the vaccination effectiveness with Pfizer-Biontech against infection is only at 35% for the Omicron VOC (5). This was corroborated by another *in vitro* study reporting an 8.4-fold reduction in neutralization for the Omicron VOC vs. the PV-D614G reference strain, whereas there was only a 1.6-fold reduction in neutralization for the Delta VOC (27). Therefore, the advantage of the Omicron VOC seems to be a combination of high transmissibility and increased immune evading abilities.

Studies on the transmission of the Omicron VOC are yet sparse and a critical prerequisite for effective control of this variant worldwide (3). In particular, it is important to clarify whether the growth advantage can be ascribed to immune evasiveness, i.e., a higher proportion of vaccinated or previously infected individuals being susceptible to infection, an increased inherent transmissibility for this variant, or both.

The aim of the present study is to investigate the household transmission of the Omicron VOC. Specifically, we address the following questions: 1) Is the secondary attack rate higher for the Omicron VOC than for the Delta VOC? 2) Does the Omicron VOC show a higher immune evasiveness relative to the Delta VOC? 3) Is booster vaccination effective for reducing transmission?

## 3 Data and Methods

### 3.1 Study design and participants

Since July 2021, the Delta VOC has been the dominant variant in Denmark. The first Danish case infected with the Omicron VOC was detected on 22^nd^ November 2021 (Danish Covid-19 Genome Consortium, DCC), and community transmission was determined to be present by late November 2021. On 8^th^ December, Danish authorities discontinued intensive contact tracing of close contacts for cases specifically infected with the Omicron VOC. We therefore started the study period on 9^th^ December 2021 when cases of both variants were treated approximately equally, thus reducing bias from intensified contact tracing and active case finding of the Omicron VOC that was implemented shortly after it’s discovery in Denmark (24). The end of the inclusion period for primary cases was set at 12^th^ December to balance the inclusion of enough cases for proper estimation and early dissemination of the results. Potential secondary cases were followed up to 7 days after, i.e., until 19^th^ December 2021 to allow for test results to be obtained. We obtained the last test results on 21^st^ December.

We used Danish register data for this study. All individuals in Denmark have a unique identification number, enabling cross linking between administrative registers. Using this, we obtained individual level information on home address, data on all antigen and RT-PCR tests for SARS-CoV-2 from the Danish Microbiology Database (MiBa; (20)), and records in the Danish Vaccination Register (13).

Households were defined based on residential addresses. We included households with 2-6 members to exclude care facilities and other places, where many individuals share the same address. If two individuals tested positive on the same day, we excluded these households from the data, to ensure proper identification of the primary case within each household.

We defined a primary case as the first individual within a household to test positive with an RT-PCR test within the study period. We followed all tests of other household members in the study period. A positive secondary case was defined by either a positive RT-PCR test or a positive antigen test (10). Almost all samples that tested positive with RT-PCR were tested with Variant PCR to determine the VOC (21) (see Appendix section 6.2). Based on the variant PCR test result of the primary case, we classified households into households with either the Omicron or the Delta VOC. The Delta VOC has been the dominating variant in Denmark since early July 2021, accounting for approximately 100% of all positive RT-PCR samples August-November 2021 (23).

We classified individuals by vaccination status into three groups: i) unvaccinated; ii) fully vaccinated (defined by the vaccine used, Comirnaty (Pfizer/BioNTech): 7 days after second dose; Vaxzevria (AstraZeneca): 15 days after second dose; Spikevax (Moderna): 14 days after second dose; Janssen (Johnson & Johnson): 14 days after vaccination, and 14 days after the second dose for cross vaccinated individuals) or 14 days after previous infection; or iii) booster-vaccinated (defined by 7 days after the booster vaccination, (16; 2)). Partially vaccinated individuals were regarded as unvaccinated in this study. By 22^nd^ December 2021, of all vaccinated individuals, 85% were vaccinated with Comirnaty, 14% with Spikevax, 1% with Janssen, and approximately 0% with AstraZeneca (22).

### 3.2 Statistical analyses

We defined the secondary attack rate (SAR) within households as the proportion of potential secondary cases within the same household that tested positive between 1-7 days following the positive test of the primary case within the household (15). Adjusted odds ratios (OR) were estimated from multivariable logistic regression models fit to the binary outcome of test status of each potential secondary case, with the primary explanatory variable reflecting the strain type in the household (Omicron vs. Delta VOC) and potentially confounding variables of age and sex of the primary case, age and sex of the potential secondary case, and household size (2-6 individuals). To test if vaccine status conferred differential protection against the Omicron and Delta VOC, we included an interaction term between vaccination status of primary and potential secondary cases and the variant. Standard errors were adjusted to control for clustering at the household level.

We have conducted a number of supplementary analyses to support the main analysis. We re-analysed each strata of the data separately using another set of logistic regression models (see appendix 7.4). To examine the potential mediating role of viral load of primary cases infected with the Omicron VOC relative to the Delta VOC, we plotted the distributions of Ct values for each variant (appendix Figure 3). We also examined the extent to which the Ct value of the primary case could explain the difference in transmission between the variants (see appendix Table 8). Our study relies on Variant PCR testing to determine if each primary case was Delta or Omicron. We estimated the intra-household correlation of variants, i.e., the probability that the positive secondary case was infected with the same variant as the primary case. To investigate if there was bias in the selection of samples for Variant PCR, we investigated the probability of sampling for Variant PCR by sample Ct value and age. Moreover, we tested the robustness of potential secondary cases being tested and testing positive by only using RT-PCR tests, which are more sensitive.

### 3.3 Ethical statement

This study was conducted using data from national registers only. According to Danish law, ethics approval is not needed for this type of research. All data management and analyses were carried out on the Danish Health Data Authority’s restricted research servers with project number FSEID-00004942. The study only contains aggregated results and no personal data. The study is, therefore, not covered by the European General Data Protection Regulation (GDPR).

### 3.4 Data availability

The data used in this study are available under restricted access due to Danish data protection legislation. The data are available for research upon reasonable request to The Danish Health Data Authority and Statens Serum Institut and within the framework of the Danish data protection legislation and any required permission from Authorities. We performed no data collection or sequencing specifically for this study.

## 4 Results

A total of 2,225 primary cases with the Omicron VOC and 9,712 primary cases with the Delta VOC were included (Table 1). The SAR was 31% in households with the Omicron VOC and 21% in households with the Delta VOC. Generally, the estimated SAR was higher for the Omicron VOC than for the Delta VOC, for all age groups. Unvaccinated potential secondary cases experienced similar attack rates in households with the Omicron VOC and the Delta VOC (29% and 28%, respectively), while fully vaccinated individuals experienced secondary attack rates of 32% in household with the Omicron VOC and 19% in households with the Delta VOC. For booster-vaccinated individuals, Omicron was associated with a SAR of 25%, while the corresponding estimate for Delta was only 11%.

**Table 1:** Summary Statistics

|  | Omicron |  |  |  | Delta |  |  |  |
| --- | --- | --- | --- | --- | --- | --- | --- | --- |
|  | Primary<br>Cases | Potential<br>Secondary<br>Cases | Positive<br>Secondary<br>Cases | SAR<br>(%) | Primary<br>Cases | Potential<br>Secondary<br>Cases | Positive<br>Secondary<br>Cases | SAR<br>(%) |
| **Total** | 2,225 | 4,718 | 1,474 | 31 | 9,712 | 23,156 | 4,923 | 21 |
| **Sex** |  |  |  |  |  |  |  |  |
| Male | 1,149 | 2,266 | 665 | 29 | 4,987 | 11,372 | 2,317 | 20 |
| Female | 1,076 | 2,452 | 809 | 33 | 4,725 | 11,784 | 2,606 | 22 |
| **Age** |  |  |  |  |  |  |  |  |
| 0-10 | 130 | 709 | 197 | 28 | 2,417 | 4,615 | 1,134 | 25 |
| 10-20 | 454 | 917 | 215 | 23 | 1,884 | 4,466 | 652 | 15 |
| 20-30 | 723 | 975 | 281 | 29 | 1,242 | 2,026 | 325 | 16 |
| 30-40 | 298 | 508 | 219 | 43 | 988 | 3,867 | 934 | 24 |
| 40-50 | 282 | 780 | 268 | 34 | 1,253 | 4,893 | 1,053 | 22 |
| 50-60 | 238 | 633 | 226 | 36 | 1,120 | 2,106 | 498 | 24 |
| 60-70 | 78 | 134 | 48 | 36 | 573 | 794 | 256 | 32 |
| 70+ | 22 | 62 | 20 | 32 | 235 | 389 | 71 | 18 |
| **Household size** |  |  |  |  |  |  |  |  |
| 2 | 865 | 865 | 362 | 42 | 2,941 | 2,941 | 833 | 28 |
| 3 | 543 | 1,086 | 341 | 31 | 2,062 | 4,124 | 866 | 21 |
| 4 | 558 | 1,674 | 489 | 29 | 3,068 | 9,204 | 1,933 | 21 |
| 5 | 202 | 808 | 229 | 28 | 1,318 | 5,272 | 1,033 | 20 |
| 6 | 57 | 285 | 53 | 19 | 323 | 1,615 | 258 | 16 |
| **Immunity** |  |  |  |  |  |  |  |  |
| Unvaccinated | 368 | 1,156 | 340 | 29 | 4,629 | 7,410 | 2,044 | 28 |
| Fully vaccinated / previous infection | 1,752 | 3,257 | 1,057 | 32 | 4,797 | 14,239 | 2,714 | 19 |
| Booster-vaccinated | 105 | 305 | 77 | 25 | 286 | 1,507 | 165 | 11 |

Notes: The secondary attack rate (SAR) is expressed as a percentage (%). Summary statistics based on primary cases are shown separately from summary statistics on potential secondary cases, positive secondary cases and SAR.

We found that the cumulative probability of potential secondary cases to be tested at least once increased from 33-41% to around 87-89% at 7 days after the primary case tested positive (Figure 1, panel a). The probability was higher when the primary case was infected with the Delta VOC compared to the Omicron VOC. The probability of potential secondary cases being tested twice increased from 9% to 69-74% at 7 days after the primary case tested positive. The probability of potential secondary cases testing positive increased from 3-4% on day 1, to 21% and 31% on day 7, when the primary case was infected with the Delta VOC and Omicron VOC, respectively (Figure 1, panel b).

**Figure 1:**
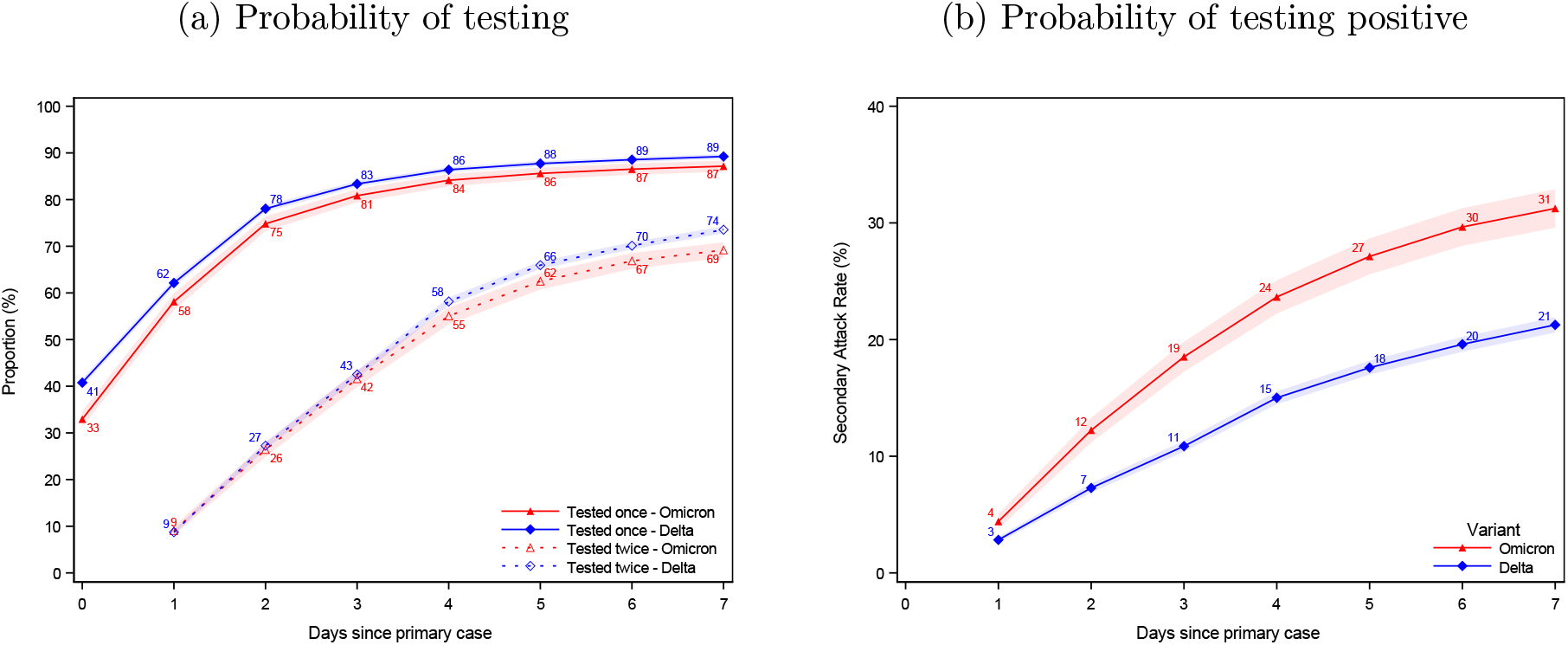
Probability of being tested and testing positive Notes: Panel (a) shows the probability of potential secondary cases being tested after a primary case has been identified within the household. Panel (b) shows the probability of potential secondary cases that test positive subsequently to a primary case being identified within the household. Note that the latter is not conditional on being tested, i.e. the denominator contains test negative individuals and untested individuals. The x axes shows the days since the primary case tested positive, and the y axes shows the proportion of individuals either being tested (a) or testing positive (b) with either antigen or RT-PCR tests, based on the variant of the primary case. The SAR for each day relative to the primary case can be read directly from panel (b). For example, the SAR on day 7 is 31% and 21%, whereas the SAR on day 4 is 24% and 15%. The shaded areas show the 95% confidence bands clustered on the household level. See appendix Figure 5 for the same two panels, only using RT-PCR tests.

The effect of vaccination on susceptibility and transmissibility of SARS-CoV-2 within households is shown in Table 2. The estimates of susceptibility by vaccine status were stratified by variant because we observed an interaction between variant and vaccination status of the potential secondary cases (p < 0.001). After adjustment for confounders, we found that in households with the Delta VOC, the OR of infection was 2.31 (95% confidence interval (CI) 2.09-2.55) for unvaccinated individuals and 0.38 (CI: 0.32-0.46) for booster-vaccinated individuals when compared to fully vaccinated potential secondary cases. For households with the Omicron VOC, the corresponding OR for infection for unvaccinated individuals was 1.04 (CI: 0.87-1.24) and 0.54 (CI: 0.40-0.71) for booster-vaccinated individuals. When considering the vaccine status of primary cases, i.e. transmissibility, we observed no difference in the OR of infection between households with the Omicron and Delta VOC. An unvaccinated primary case was associated with an OR of 1.41 (CI: 1.27-1.57) for potential secondary cases compared to fully vaccinated primary cases, while a booster-vaccinated primary case was associated with a decreased OR of 0.72 (CI: 0.56-0.92).

**Table 2:** Effect of Vaccination

|  | Susceptibility |  | Transmissibility |
| --- | --- | --- | --- |
|  | Omicron households | Delta households | All households |
| Unvaccinated | 1.04<br>(0.87-1.24) | 2.31<br>(2.09-2.55) | 1.41<br>(1.27-1.57) |
| Fully vaccinated | ref<br>(.) | ref<br>(.) | ref<br>(.) |
| Booster-vaccinated | 0.54<br>(0.40-0.71) | 0.38<br>(0.32-0.46) | 0.72<br>(0.56-0.92) |
| Number of observations | 27,874 | 27,874 | 27,874 |
| Number of households | 11,937 | 11,937 | 11,937 |

The relative difference in transmission between the Omicron and Delta variants when comparing potential secondary cases with the same vaccine status is shown in Table 3. For unvaccinated individuals, an OR of 1.17 (CI: 0.99-1.38) was estimated for households with the Omicron VOC compared to households with the Delta VOC. For fully vaccinated individuals, the OR was 2.61 (CI: 2.34-2.90) for households with the Omicron VOC, and for booster-vaccinated individuals the OR was 3.66 (CI: 2.65-5.05). This significant change in OR for fully vaccinated and booster-vaccinated individuals represents strong evidence of immune evasion of the Omicron VOC.

**Table 3:**
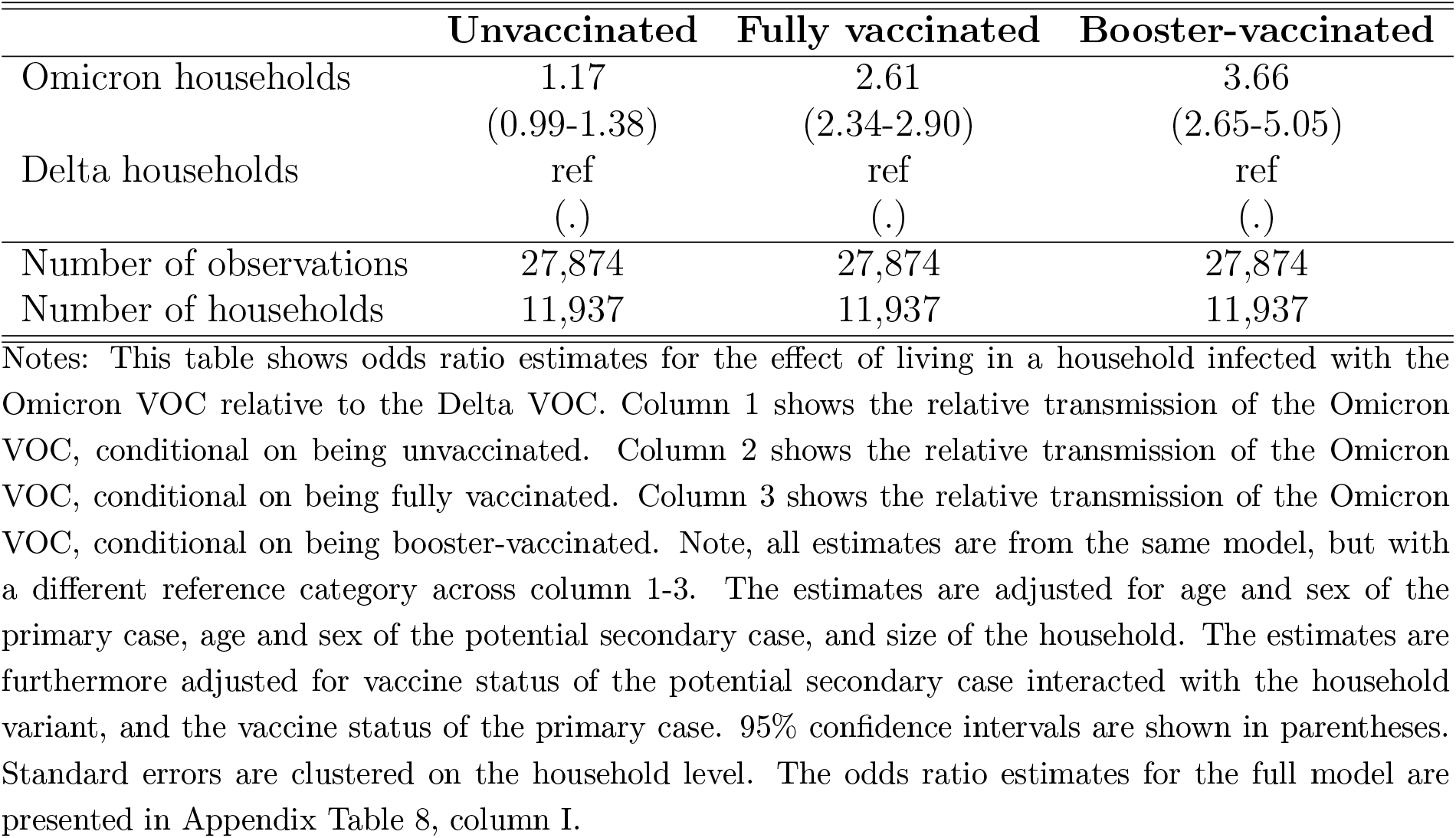
Relative effect of the Omicron VOC

|  | **Unvaccinated** | **Fully vaccinated** | **Booster-vaccinated** |
| --- | --- | --- | --- |
| Omicron households | 1.17<br>(0.99-1.38) | 2.61<br>(2.34-2.90) | 3.66<br>(2.65-5.05) |
| Delta households | ref<br>(.) | ref<br>(.) | ref<br>(.) |
| Number of observations | 27,874 | 27,874 | 27,874 |
| Number of households | 11,937 | 11,937 | 11,937 |

We compared the Ct values of primary cases with the Omicron VOC and the Delta VOC (Appendix 7, Figure 3). The distribution of Ct values for cases with the Omicron VOC were slightly skewed to the left compared to cases with the Delta VOC, but the median values (27.24 and 28.29, respectively) did not differ substantially. Adjustment for Ct values of the primary cases did not materially alter the findings, suggesting that the difference between the Omicron and Delta VOC transmission is not due to differences in viral load in the primary case (appendix Table 8). Similarly, the distribution of time since last vaccination/booster/infection among positive secondary cases were approximately the same across the two variants (appendix Figure 4). However, these analyses are limited by the fact that vaccine roll out in Denmark is largely determined by age, which makes it difficult to estimate true associations for variables that are also correlated with age.

**Figure 2:**
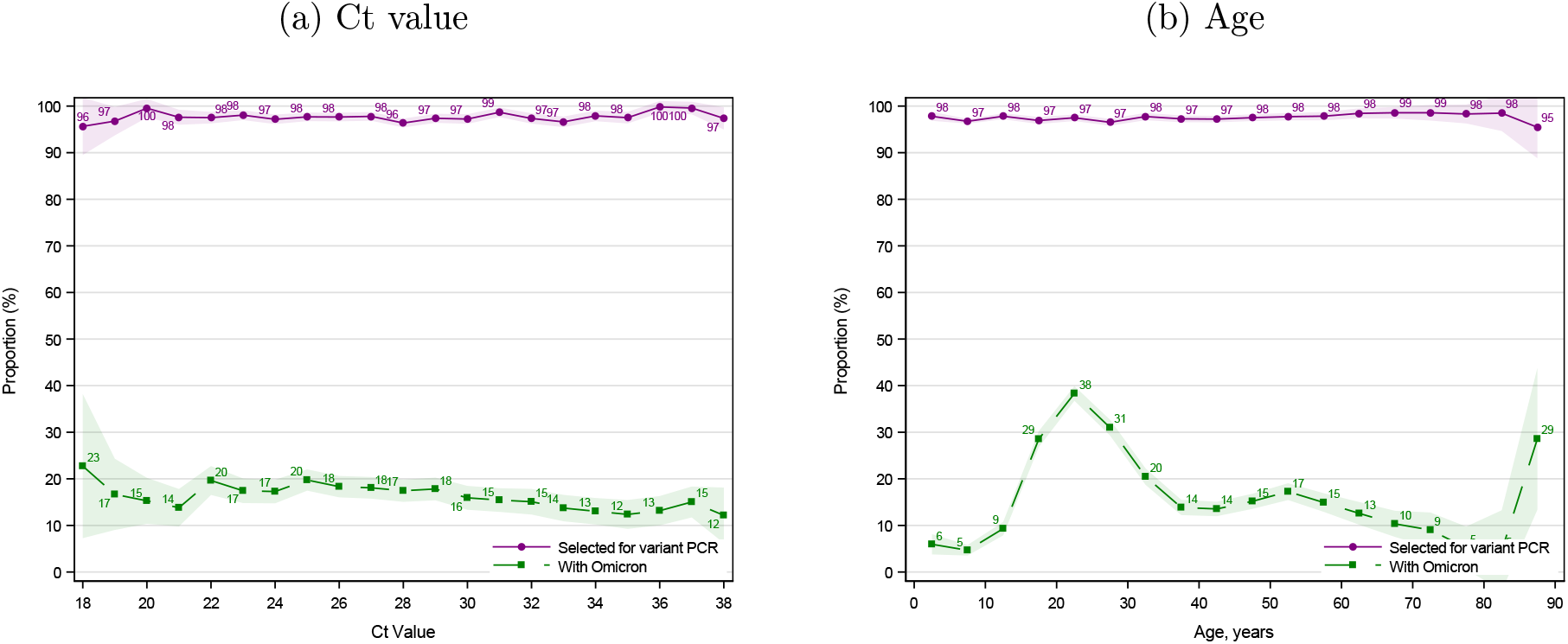
Probability of sampling for Variant PCR Notes: This figure shows the proportion of positive RT-PCR samples selected (purple) for Variant PCR testing and the proportion testing positive (green) with the Omicron VOC. Panel (a) shows the selection by Ct value; panel (b) by age. Only positive RT-PCR tests from 9^th^-12^th^ December 2021 performed by TestCenter Denmark are included. The shaded areas show the 95% confidence bands.

**Figure 3:**
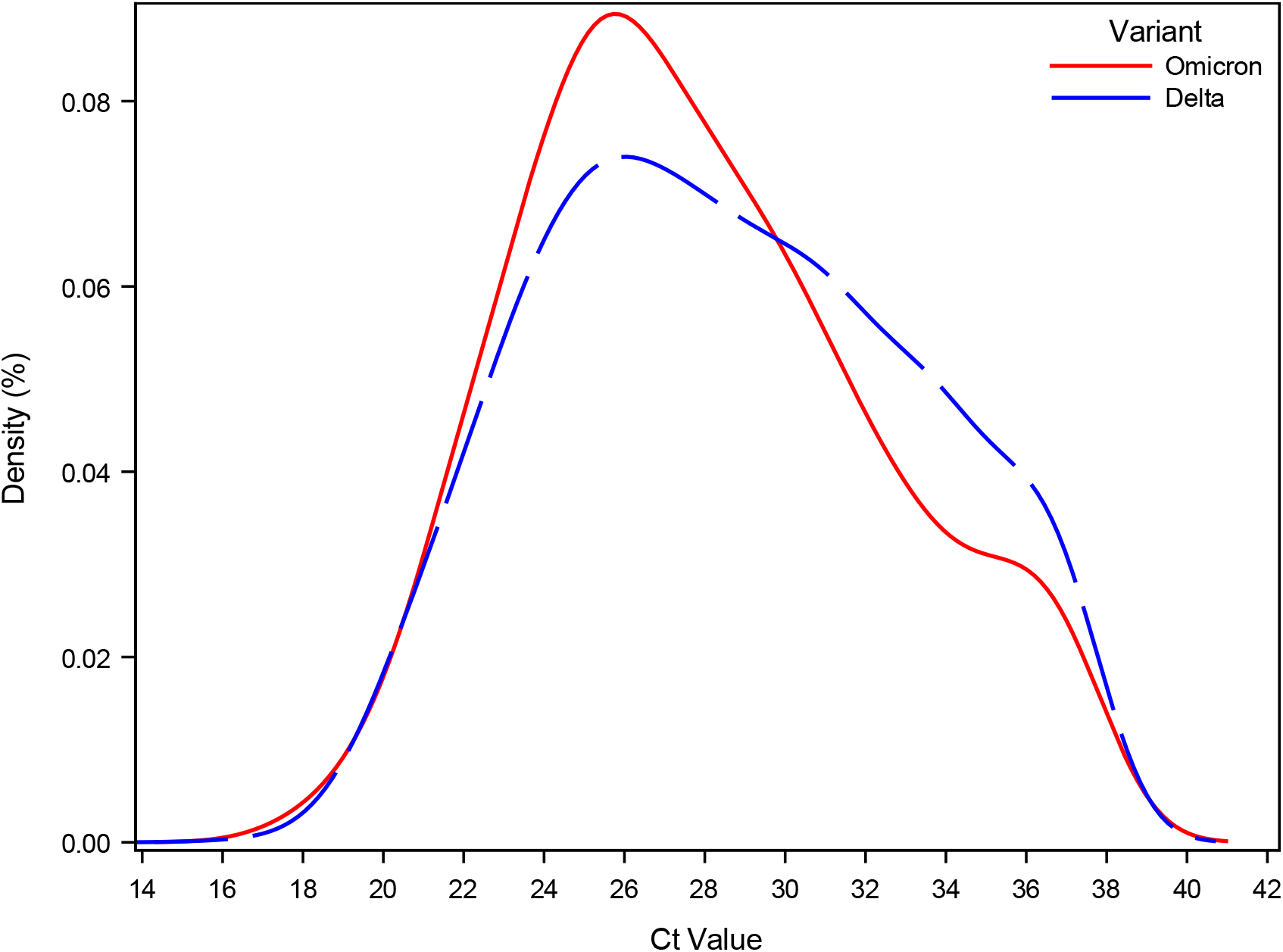
Density of Ct value Notes: This figure shows the density of the sample Ct values of primary cases stratified by the Omicron and Delta VOC.

**Figure 4:**
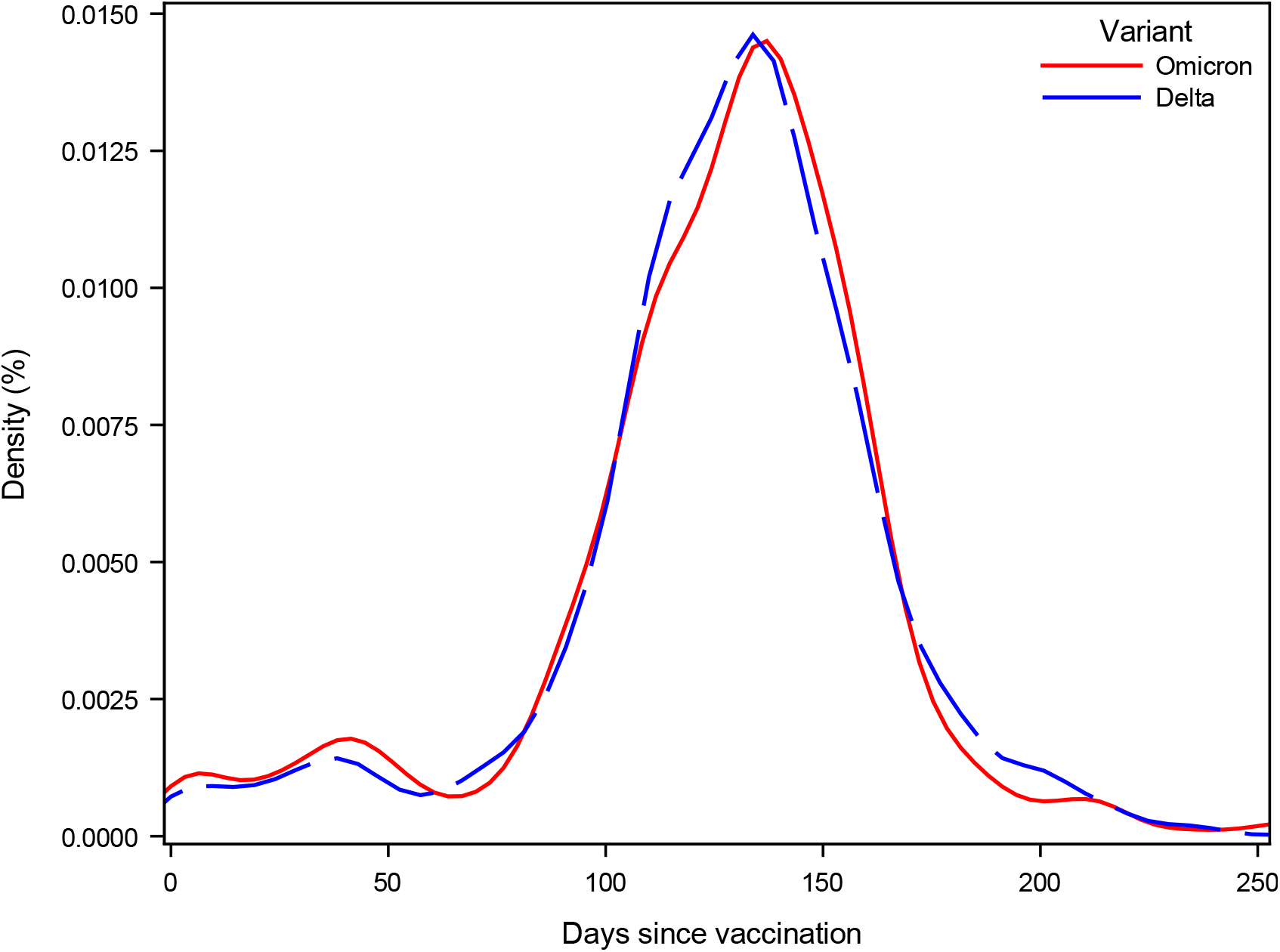
Time since vaccination for positive secondary cases Notes: This figure shows the distribution of days since last vaccination/infection for positive secondary cases, stratified by the household VOC.

The probability that a sample was selected for Variant PCR was stable across Ct value, varying between 97% and 100% (Appendix 7, Figure 2). Selection for Variant PCR was also similar across age groups, varying between 95% and 99%. The Variant PCR was validated with high quality whole genome sequencing (WGS) data, and showed an estimated false negative rate of 0.06% and a false positive rate of 1.15% (Appendix section 6.3).

We found an intra-household correlation between primary case and secondary case variants of 98.0% and 97.7% for households with primary cases infected with the Omicron VOC and the Delta VOC, respectively (Appendix section 7.2). This supports the assumption that subsequent infections within the same household are likely to be associated with the same variant as was identified for the primary case.

Finally, misclassification of primary and secondary cases is also a potential concern. It is not given that the index case is actually also the case with the first positive RT-PCR test. However, we believe that this potential bias would impact both the Delta and Omicron VOC transmission estimates equally. Figure 1, panel b, shows the proportion of secondary cases testing positive on each day after the primary case. One day after, 3% and 4% have tested positive, two days after, 7% and 12% have tested positive. These cases could have been misclassified as secondary cases, instead of primary cases. To investigate the sensitivity of our results for this potential misclassification, we estimated our regression model including only secondary cases that tested positive on day 2-7 and on day 3-7 (Appendix Table 9).

## 5 Discussion and Conclusion

Our results show that the Omicron VOC is generally 2.7-3.7 times more infectious than the Delta VOC among vaccinated individuals (Table 3). This observation is in line with data from (18), which estimated that 19% of Omicron VOC primary cases in households in the UK resulted in at least one other infection within the household, compared to only 8.3% of those associated with the Delta VOC. Furthermore, we show that fully vaccinated and booster-vaccinated individuals are generally less susceptible to infection compared to unvaccinated individuals (Table 2). We also show that booster-vaccinated individuals generally had a reduced transmissibility (OR: 0.72, CI: 0.56-0.92), and that unvaccinated individuals had a higher transmissibility (OR: 1.41, CI: 1.27-1.57), compared to fully vaccinated individuals.

Surprisingly, we observed no significant difference between the SAR of Omicron versus Delta among unvaccinated individuals (Table 3). This indicates that the increased transmissibility of the Omicron VOC primarily can be ascribed to immune evasion rather than an inherent increase in the basic transmissibility. If this observation can be confirmed by independent studies, it has important ramifications for the understanding of the current challenges for control of the epidemic. Our data indicate that the non-pharmaceutical interventions that were used to control the previous variants of SARS-CoV-2 are also likely to be effective against the Omicron VOC. On the other hand, although we showed that booster vaccines did offer some protection against household transmission, the reduced level of protection means that vaccination is less likely to be sufficient to curb transmission within a population. Furthermore, the duration of the protective effect is currently unknown, and the rapidly waning effectiveness of the second dose against the Omicron VOC as well as data from neutralization assays (14; 8) do raise some concerns about the longevity of the booster response. This means that the current vaccines are unlikely to mitigate the spread of the Omicron VOC to the extent that has been achieved for previous variants on the long term. We therefore suggest that adapted or improved vaccines may be necessary to mitigate the spread of the Omicron VOC. However, both a primary series and a booster dose is likely to play an important role in reducing transmission on a short term and modifying the outcome of infection by reducing severity. We were unable to address this question as detailed clinical data were not available in our registries, and in any case the relatively short follow-up time precludes a thorough investigation of any subsequent hospitalisations and deaths.

(1) note that the existing circulating immunity within a country is of major importance in limiting the severity of the epidemic with the Omicron VOC. We found that booster-vaccinated individuals in Omicron VOC households had an OR of 0.54 (CI: 0.40-0.71) compared to fully vaccinated individuals (Table 2). This shows that booster vaccination is effective for reducing household transmission of the Omicron VOC. However, this effect is also affected by other factors such as waning immunity for fully vaccinated individuals, since they were typically vaccinated much earlier compared to the dose received as a booster vaccination. For the Delta VOC, the booster vaccination had an OR of 0.38 (CI: 0.32-0.46) compared to fully vaccinated individuals, showing the high effectiveness of booster vaccinations against the Delta VOC. These estimates are important for decision makers worldwide, as they can inform models for predicting the epidemic and thus for balancing the most appropriate level of restrictions to control transmission in different situations.

There are some potential biases in this study. Firstly, in this initial phase, the spread of the Omicron VOC was characterized by spreading events, and has therefore not yet reached an even distribution throughout the population (3). We found that a large proportion of the Omicron VOC cases were aged 20-30 years, and therefore different from the age distribution of Delta cases (Table 1). Furthermore, the Omicron VOC is mainly found in households with 2 members, whereas the Delta VOC is mainly found in households with 4 members. This could potentially impact the comparability of our estimates for the two the variants. However, our results are robust, when we exclude households with a primary case <10 years and when we only include 2-person households (Appendix Table 8).

Recently, self testing kits have become widely available for purchase in Denmark. This could influence the results, for instance if individuals that self-test at home refrain from also being tested in public testing facilities, meaning that their test results are not registered in the national databases. However, the probability of being tested was very similar between RT-PCR and any test (Figure 1 and Appendix Figure 5), and consequently most positive antigen tests were confirmed with a positive RT-PCR test, so we assume that has likely also been the case for positive self-tests as well. Moreover, we believe that this potential bias will impact both the Delta and the Omicron VOC transmission estimates equally.

**Figure 5:**
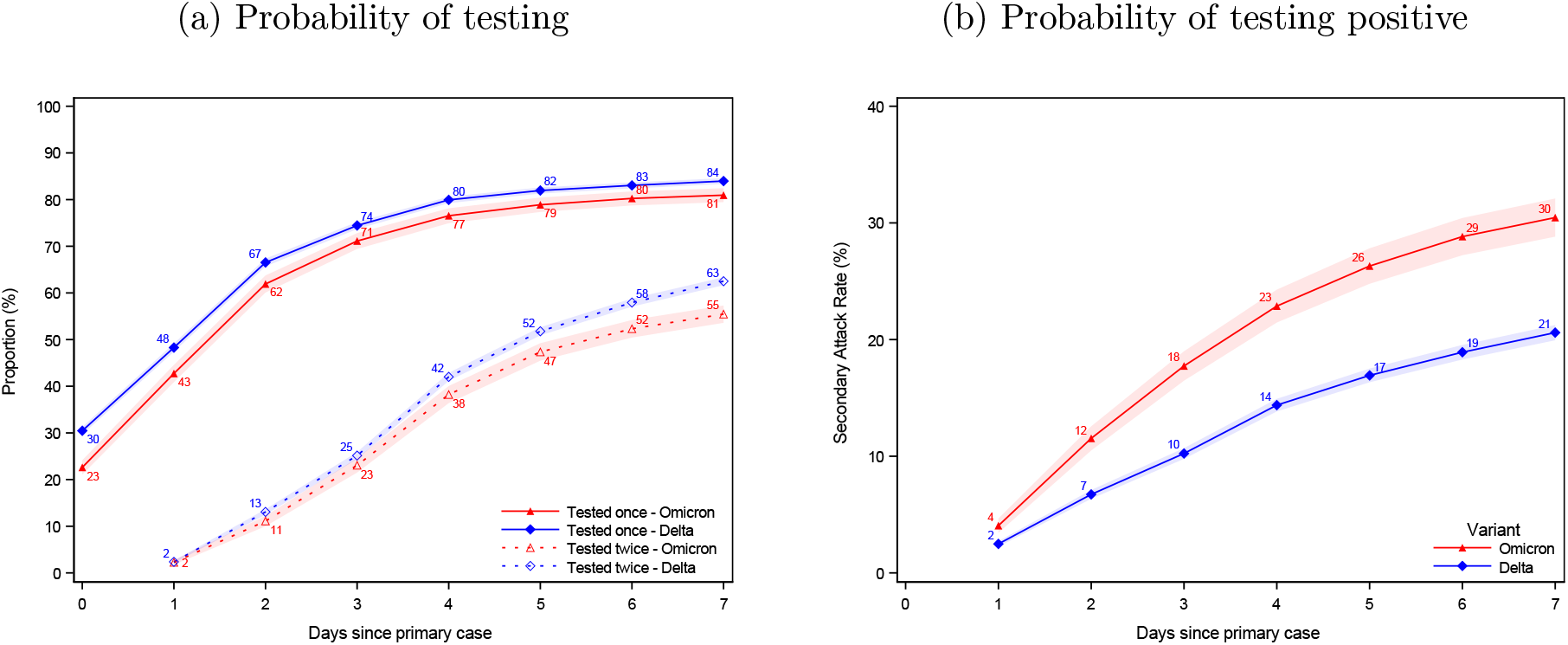
Probability of being tested and testing positive with an RT-PCR test Notes: This figure shows the same as Figure 1, but only including RT-PCR tests. Panel (a) shows the probability of potential secondary cases being tested after a primary case has been identified within the household. Panel (b) shows the probability of potential secondary cases that test positive subsequently to a primary case being identified within the household. Note that the latter is not conditional on being tested, i.e. the denominator contains test negative individuals and untested individuals. The x axes shows the days since the primary case tested positive, and the y axes shows the proportion of individuals either being tested (a) or testing positive (b) with an RT-PCR test, based on the variant of the primary case. The SAR for each day relative to the primary case can be read directly from panel (b). For example, the SAR on day 7 is 30% and 21%, whereas the SAR on day 4 is 23% and 14%. The shaded areas show the 95% confidence bands clustered on the household level.

There are also a number of other confounding variables that might lead to biases within our study. Vaccines are not randomly distributed within the population, so immunocom-promised and other vulnerable individuals are more likely to have had access to a booster vaccine in particular. To some extent, this has been addressed by including age as an explanatory variable in the models, but it was not possible to include information on underlying risk conditions, so this information is incomplete. Similarly, there are likely underlying behavioural drivers for an individual being unvaccinated, which are likely to confound with other risky behaviours that might be expected to increase both transmission and susceptibility to infection (e.g. poor use of face masks, reduced attention to hygiene). The use of registry data limits our inference to associations between transmission/susceptibility and vaccination status of individuals, where part of the association is due to general characteristics of the individuals themselves rather than their vaccination status. However, a key point of our study is that these biases are non-differential with respect to variant, i.e. they would be expected to affect transmission of both the Delta and Omicron VOC identically because risky behaviours and/or underlying health concerns should affect both variants equally. Consequently, although the associations between vaccination status and transmission for each variant are likely to each be susceptible to bias, we believe that any such bias is non-differential with respect to variant. Therefore, our results concerning the relatively higher transmissibility of the Omicron vs. Delta VOC for vaccinated individuals should be robust to these potential biases.

The SAR was found to be higher for the Omicron VOC than for the Delta VOC across all age groups (Table 1). Notably, the SAR for individuals aged 70+ years was 32% for the Omicron VOC compared to only 18% for the Delta VOC. This has implications for e.g. care facilities, highlighting the need for increased protection against transmission, now that the Omicron VOC will likely overtake the Delta VOC in many countries.

To conclude, we found an increased susceptibility for unvaccinated individuals, and a reduced susceptibility for booster-vaccinated individuals, compared to fully vaccinated individuals in households infected with the Delta VOC. Additionally, we found a reduced susceptibility for booster-vaccinated individuals in households infected with the Omicron VOC. Furthermore, we found an increased transmissibility from unvaccinated individuals, and a reduced transmissibility from booster-vaccinated individuals, compared to fully vaccinated individuals. Lastly, we we found a general higher transmission in households infected with the Omicron VOC relative to the Delta for both unvaccinated, fully vaccinated and booster-vaccinated individuals. The Omicron VOC showed immune evasiveness for fully vaccinated and booster-vaccinated individuals. Our results confirm that booster vaccination has the potential to reduce Omicron VOC transmission in households, although vaccination as a strategy for epidemic control is increasingly challenged by the immune evasiveness of the Omicron VOC.

Comparing Omicron VOC to Delta VOC, we found an 1.17 (95%-CI: 0.99-1.38) times higher SAR for unvaccinated, 2.61 times (95%-CI: 2.34-2.90) higher for fully vaccinated and 3.66 (95%-CI: 2.65-5.05) times higher for booster-vaccinated individuals, demonstrating strong evidence of immune evasiveness of the Omicron VOC.

## Aknowledgements

We thank Statens Serum Institut and The Danish Health Data Authority for collecting and providing access to data access. We also thank the rest of the Expert Group for Mathematical Modelling of COVID-19 at Statens Serum Institut for helpful discussions. The authors wish to thank the Danish Covid-19 Genome Consortium for genotyping SARS-CoV-2 positive samples.

## Contributions

FPL performed all data analyses. LEC, MD, FPL, LHM and CTK designed the study and devised the statistical analysis. FPL, CTK and LHM wrote the first draft. All other authors contributed to the discussion, revised the first draft and approved the submitted version.

## Supplementary Appendix

### 6 Background

This section provides some background information in order to better understand the circumstances underlying our study. The results build on all data available in Denmark, i.e., not restricted to our study sample.

#### 6.1 Time to test result

Denmark provides free testing with both antigen and RT-PCR tests. Antigen tests provide a quick test result (positive/negative) with a median time from sample to result of less than 30 minutes (Table 4). All positive antigen tests are recommended to be confirmed with an RT-PCR test. RT-PCR tests are more sensitive (25), but also require a longer time before the result is known. The median time is approximately 24 hours (Table 5). Only samples from positive RT-PCR tests are selected for Variant PCR and whole genome sequencing.

**Table 4:**
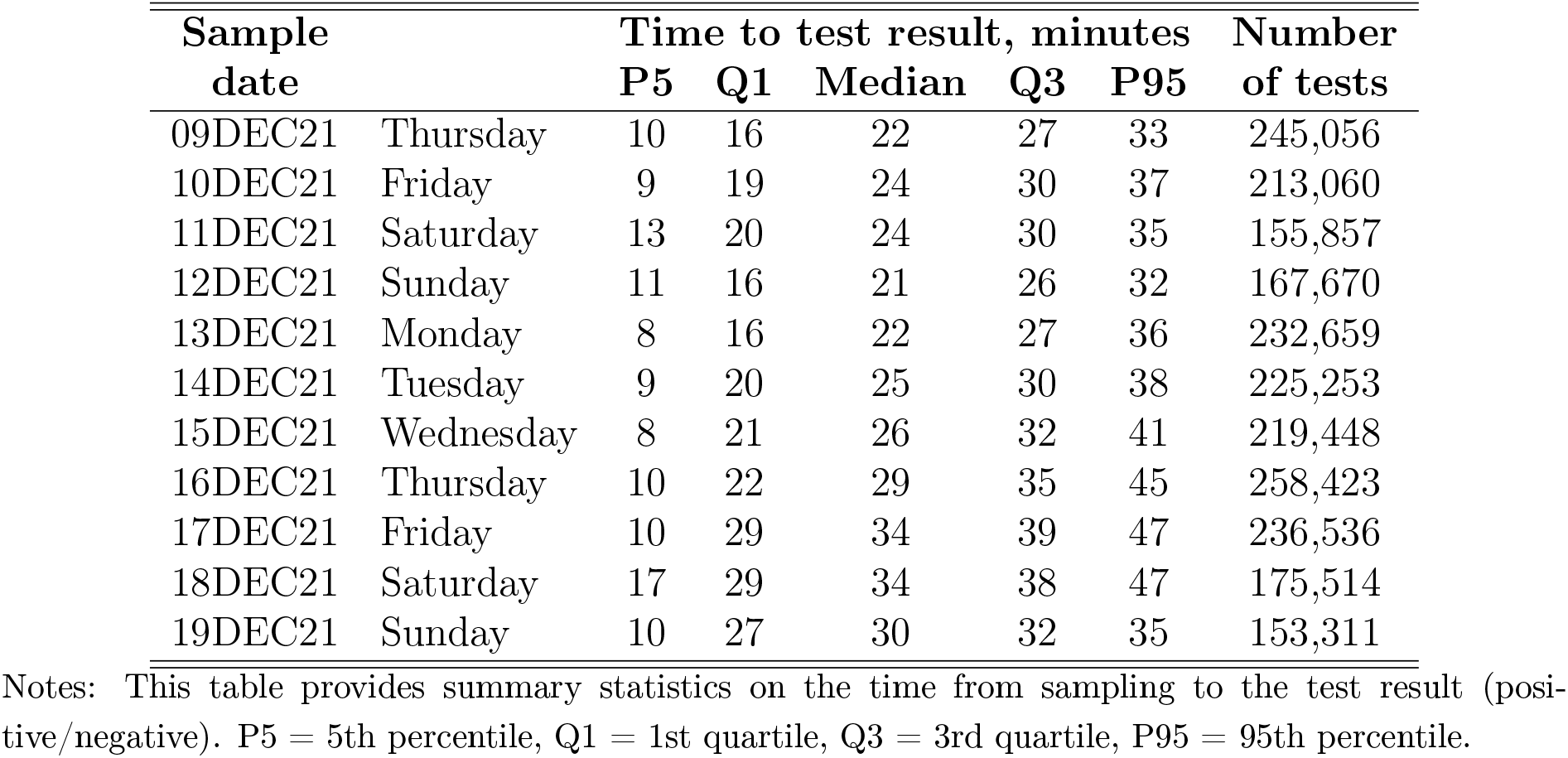
Time to test results, antigen tests, minutes

**Table 5:** Time to test results, RT-PCR tests, hours

| Sample date |  | Time to test result, hours |  |  |  |  | Number of PCR tests |
| --- | --- | --- | --- | --- | --- | --- | --- |
|  |  | P5 | Q1 | Median | Q3 | P95 |  |
| 09DEC21 | Thursday | 16 | 20 | 22 | 28 | 41 | 268,357 |
| 10DEC21 | Friday | 16 | 20 | 23 | 28 | 41 | 274,827 |
| 11DEC21 | Saturday | 16 | 20 | 24 | 29 | 40 | 178,176 |
| 12DEC21 | Sunday | 16 | 20 | 24 | 28 | 37 | 178,066 |
| 13DEC21 | Monday | 16 | 20 | 23 | 28 | 41 | 261,215 |
| 14DEC21 | Tuesday | 16 | 20 | 22 | 27 | 40 | 254,107 |
| 15DEC21 | Wednesday | 16 | 20 | 24 | 29 | 42 | 224,897 |
| 16DEC21 | Thursday | 16 | 20 | 24 | 29 | 42 | 255,183 |
| 17DEC21 | Friday | 16 | 20 | 24 | 29 | 45 | 273,041 |
| 18DEC21 | Saturday | 16 | 21 | 25 | 29 | 42 | 221,572 |
| 19DEC21 | Sunday | 16 | 20 | 24 | 29 | 39 | 220,871 |

Notes: This table provides summary statistics on the time from sampling to the test result (positive/negative). P5 = 5th percentile, Q1 = 1st quartile, Q3 = 3rd quartile, P95 = 95th percentile.

#### 6.2 Probability of sampling for Variant PCR

This subsection provides details on the sampling probability for variant PCR.

Figure 2 shows the proportion of positive RT-PCR samples selected for Variant PCR testing and the proportion testing positive with the Omicron VOC. There was no clear selection bias in the probability that a positive RT-PCR sample was selected for Variant PCR testing (purple)—neither across sample Ct value (panel a) nor across age (panel b). There was a higher proportion of cases aged 15-30 years that tested positive with the Omicron VOC (green, panel b).

#### 6.3 Robustness of Variant PCR results

This subsection provides additional results in order to examine the robustness of the Variant PCR used to detect the Omicron VOC.

The Variant PCR was validated using the available high quality whole-genome sequencing (WGS) data. The rate of false positives was estimated to 1.15% (18/1,567) (Table 6, panel I). The rate of false negative PCR results was estimated to 0.06% (1/1,557) (Table 6, panel II).

**Table 6:** Validation of Variant PCR test using whole genome sequencing (WGS).

| **I. False positive rate** |  |  |
| --- | --- | --- |
|  | **Number of samples** | **%** |
| **Confirmed by WGS** |  |  |
| No (false positive rate) | 18 | 1.15 |
| Yes | 1,549 | 98.85 |
| Total | 1,567 | 100.00 |
| **II. False negative rate** |  |  |
|  | **Number of samples** | **%** |
| **Variant PCR result** |  |  |
| Omicron | 1,549 | 99.49 |
| Inconclusive | 7 | 0.45 |
| Delta (false negative rate) | 1 | 0.06 |
| Total | 1,557 | 100.00 |

Notes: This table shows the estimates of the false positive rate and the false negative rate of the Variant PCR test, using whole genome sequencing (WGS). Panel I shows the number of Variant PCR tests that were confirmed (Yes) or not confirmed (No) using high quality whole-genome sequencing (WGS). The estimated false positive rate was 1.15% (18/1,567). Panel II shows the number of Variant PCR tests in samples confirmed as the Omicron VOC using high quality whole-genome sequencing (WGS). The estimated false negative rate was 0.06% (1/1,557).

### 7 Additional analyses

This section provides additional analyses in order to better understand our main results.

#### 7.1 Viral load of primary cases

Figure 3 shows the density of sample Ct values of primary cases stratified by the Omicron VOC and Delta VOC.

#### 7.2 Intra-household correlation of variants

In this subsection, we investigate the intra-household correlation of variants, i.e., the probability that the primary and positive secondary cases are infected with the same variant.

In households where the primary case was infected with the Omicron VOC, we found 1,160 positive secondary cases that also had a Variant PCR result (Table 7). Of these, 1,137 (98%) were also Omicron VOC and 23 (2%) were Delta VOC. Similarly, in households where the primary case was infected with the Delta VOC, we found 3,917 positive secondary cases. Of these, 3,827 (98%) were also Delta and 90 (2%) were Omicron VOC. The overall intra-household correlation of variants was 97.8 (CI: 97.3-98.3).

**Table 7:** Intra-household correlation of variants

| **I. Number of cases** |  |  |  |
| --- | --- | --- | --- |
|  | **Primary case** |  |  |
|  | **Omicron** | **Delta** | **All** |
| **Secondary Case** |  |  |  |
| Omicron | 1,137 | 90 | 1,227 |
| Delta | 23 | 3,827 | 3,850 |
| All | 1,160 | 3,917 | 5,077 |
| **II. Regression estimates** |  |  |  |
|  | **Primary case** |  |  |
|  | **Omicron** | **Delta** | **All** |
| Intra-household correlation (%) | 98.0 | 97.7 | 97.8 |
| (95%-CI) | (97.2-98.9) | (97.8-99.4) | (97.3-98.3) |
| Number of observations | 1,160 | 3,917 | 5,077 |
| Number of households | 864 | 2,866 | 3,730 |

Notes: This table provides estimates of the intra-household correlation of variants, i.e., the probability that the primary and positive secondary cases are infected with the same variant. Panel I provides the number of observations. Panel II provides regression estimates. Standard errors are clustered on the household level.

#### 7.3 Time since vaccination for positive secondary cases

Figure 4 shows the distribution of days since last vaccination/infection for positive secondary cases, stratified by the household VOC. We saw no clear evidence of difference across the two distributions.

#### 7.4 Robustness of main results

This subsection provides additional results in order to validate the robustness of the results shown in the main paper.

Figure 5 shows the same as Figure 1, but only using RT-PCR tests (i.e. excluding antigen tests).

Tables 8 and 9 show the results of logistic regression models fit to specific strata of the data.

**Table 8:** Effect of Vaccination

|  | I |  | II |  | III |  | IV |  |
| --- | --- | --- | --- | --- | --- | --- | --- | --- |
|  | OR | 95%-CI | OR | 95%-CI | OR | 95%-CI | OR | 95%-CI |
| **Potential secondary case vaccination status** |  |  |  |  |  |  |  |  |
| *Delta households* |  |  |  |  |  |  |  |  |
| Unvaccinated | 2.32 | (2.10-2.56) | 2.27 | (2.02-2.56) | 2.01 | (1.54-2.62) | 2.44 | (2.12-2.81) |
| Fully vaccinated | ref | (.) | ref | (.) | ref | (.) | ref | (.) |
| Booster-vaccinated | 0.38 | (0.32-0.45) | 0.40 | (0.33-0.50) | 0.35 | (0.26-0.47) | 0.40 | (0.32-0.52) |
| *Omicron households* |  |  |  |  |  |  |  |  |
| Unvaccinated | 2.73 | (2.29-3.26) | 2.59 | (2.14-3.13) | 2.45 | (1.50-4.00) | 3.16 | (2.47-4.05) |
| Fully vaccinated | 2.61 | (2.34-2.90) | 2.55 | (2.28-2.86) | 2.61 | (2.13-3.19) | 2.66 | (2.29-3.09) |
| Booster-vaccinated | 1.39 | (1.04-1.84) | 1.33 | (0.99-1.79) | 1.50 | (0.95-2.38) | 1.11 | (0.74-1.67) |
| **Primary case vaccination status** |  |  |  |  |  |  |  |  |
| Unvaccinated | 1.41 | (1.27-1.57) | 1.43 | (1.29-1.59) | 1.23 | (0.97-1.54) | 1.21 | (1.05-1.41) |
| Fully vaccinated | ref | (.) | ref | (.) | ref | (.) | ref | (.) |
| Booster-vaccinated | 0.72 | (0.56-0.92) | 0.71 | (0.56-0.91) | 0.72 | (0.52-1.01) | 0.58 | (0.41-0.83) |
| **Primary case age** |  |  |  |  |  |  |  |  |
| 0-10 | 1.28 | (1.10-1.50) | - | - | 0.89 | (0.56-1.43) | 1.52 | (1.23-1.88) |
| 10-20 | 1.04 | (0.90-1.21) | 1.09 | (0.94-1.26) | 1.00 | (0.70-1.43) | 1.01 | (0.83-1.23) |
| 20-30 | ref | (.) | ref | (.) | ref | (.) | ref | (.) |
| 30-40 | 1.92 | (1.65-2.25) | 1.95 | (1.66-2.28) | 1.66 | (1.21-2.27) | 1.65 | (1.33-2.04) |
| 40-50 | 2.31 | (1.98-2.70) | 2.39 | (2.05-2.79) | 1.96 | (1.41-2.75) | 2.25 | (1.83-2.77) |
| 50-60 | 2.45 | (2.09-2.87) | 2.49 | (2.13-2.92) | 2.25 | (1.69-2.98) | 2.43 | (1.96-3.02) |
| 60-70 | 3.16 | (2.54-3.94) | 3.11 | (2.50-3.88) | 2.68 | (1.92-3.74) | 2.69 | (1.97-3.68) |
| 70+ | 3.99 | (2.76-5.75) | 3.85 | (2.67-5.55) | 4.11 | (2.53-6.68) | 4.00 | (2.36-6.77) |
| **Potential secondary case age** |  |  |  |  |  |  |  |  |
| 0-10 | 0.82 | (0.71-0.95) | 0.83 | (0.71-0.98) | 0.38 | (0.22-0.66) | 0.81 | (0.66-0.99) |
| 10-20 | 0.70 | (0.61-0.80) | 0.66 | (0.57-0.77) | 0.50 | (0.34-0.74) | 0.72 | (0.60-0.87) |
| 20-30 | ref | (.) | ref | (.) | ref | (.) | ref | (.) |
| 30-40 | 1.64 | (1.44-1.88) | 1.54 | (1.31-1.79) | 0.86 | (0.63-1.17) | 1.77 | (1.46-2.13) |
| 40-50 | 1.60 | (1.40-1.82) | 1.43 | (1.24-1.65) | 0.79 | (0.56-1.11) | 1.74 | (1.44-2.09) |
| 50-60 | 1.48 | (1.29-1.71) | 1.42 | (1.23-1.65) | 1.45 | (1.10-1.92) | 1.55 | (1.28-1.89) |
| 60-70 | 1.53 | (1.25-1.87) | 1.49 | (1.21-1.83) | 1.64 | (1.18-2.28) | 1.73 | (1.32-2.28) |
| 70+ | 1.01 | (0.75-1.37) | 0.98 | (0.72-1.35) | 0.88 | (0.56-1.38) | 0.92 | (0.60-1.42) |
| **Household size** |  |  |  |  |  |  |  |  |
| 2 | 2.26 | (1.85-2.75) | 2.36 | (1.86-3.00) | - | - | 2.34 | (1.79-3.05) |
| 3 | 1.53 | (1.26-1.86) | 1.50 | (1.19-1.90) | - | - | 1.51 | (1.16-1.95) |
| 4 | 1.51 | (1.25-1.82) | 1.53 | (1.21-1.94) | - | - | 1.45 | (1.13-1.87) |
| 5 | 1.40 | (1.15-1.70) | 1.38 | (1.08-1.78) | - | - | 1.42 | (1.09-1.86) |
| 6 | ref | (.) | ref | (.) | - | - | ref | (.) |
| **Primary case sex** |  |  |  |  |  |  |  |  |
| Male | ref | (.) | ref | (.) | ref | (.) | ref | (.) |
| Female | 0.99 | (0.92-1.07) | 1.03 | (0.95-1.13) | 1.08 | (0.91-1.29) | 0.97 | (0.88-1.08) |
| **Potential secondary case sex** |  |  |  |  |  |  |  |  |
| Male | ref | (.) | ref | (.) | ref | (.) | ref | (.) |
| Female | 1.14 | (1.07-1.20) | 1.15 | (1.07-1.22) | 1.33 | (1.11-1.58) | 1.10 | (1.01-1.19) |
| **Ct value** |  |  |  |  |  |  |  |  |
| 14-16 | - | - | - | - | - | - | 1.53 | (0.30-7.78) |
| 16-18 | - | - | - | - | - | - | 3.91 | (1.75-8.71) |
| 18-20 | - | - | - | - | - | - | 3.16 | (2.19-4.56) |
| 20-22 | - | - | - | - | - | - | 2.83 | (2.09-3.83) |
| 22-24 | - | - | - | - | - | - | 2.12 | (1.60-2.80) |
| 24-26 | - | - | - | - | - | - | 1.91 | (1.45-2.52) |
| 26-28 | - | - | - | - | - | - | 1.59 | (1.20-2.10) |
| 28-30 | - | - | - | - | - | - | 1.58 | (1.19-2.10) |
| 30-32 | - | - | - | - | - | - | 1.28 | (0.96-1.71) |
| 32-34 | - | - | - | - | - | - | 1.31 | (0.98-1.77) |
| 34-36 | - | - | - | - | - | - | 1.48 | (1.09-2.02) |
| 36-38 | - | - | - | - | - | - | ref | (.) |
| Number of observations | 27,874 |  | 20,228 |  | 3,806 |  | 14,657 |  |
| Number of households | 11,937 |  | 9,390 |  | 3,806 |  | 6,242 |  |

Notes: Column I shows the odds ratio estimates of our main specification, i.e., the same as used in Table 2 and 3. Column II shows the odds ratio estimates, when excluding households with a primary case <10 years. Column III shows the odds ratio estimates, when only including 2-person households. Column IV shows the odds ratio estimates, while adjusting for Ct value of the primary case. 95%-Confidence interval in parentheses. Standard errors are clustered on the household level.

**Table 9:** Effect of Vaccination, sensitivity for days for including positive secondary cases

| Days including positive secondary cases | I |  | II |  | III |  |
| --- | --- | --- | --- | --- | --- | --- |
|  | 1-7 |  | 2-7 |  | 3-7 |  |
|  | OR | 95%-CI | OR | 95%-CI | OR | 95%-CI |
| **Potential secondary case vaccination status** |  |  |  |  |  |  |
| *Delta households* |  |  |  |  |  |  |
| Unvaccinated | 2.32 | (2.10-2.56) | 2.20 | (1.99-2.45) | 2.13 | (1.90-2.38) |
| Fully vaccinated | ref | (.) | ref | (.) | ref | (.) |
| Booster-vaccinated | 0.38 | (0.32-0.45) | 0.39 | (0.33-0.47) | 0.41 | (0.34-0.51) |
| *Omicron households* |  |  |  |  |  |  |
| Unvaccinated | 2.73 | (2.29-3.26) | 2.80 | (2.34-3.35) | 2.65 | (2.17-3.24) |
| Fully vaccinated | 2.61 | (2.34-2.90) | 2.61 | (2.33-2.92) | 2.45 | (2.17-2.78) |
| Booster-vaccinated | 1.39 | (1.04-1.84) | 1.40 | (1.04-1.89) | 1.19 | (0.83-1.71) |
| **Primary case vaccination status** |  |  |  |  |  |  |
| Unvaccinated | 1.41 | (1.27-1.57) | 1.43 | (1.28-1.59) | 1.38 | (1.23-1.55) |
| Fully vaccinated | ref | (.) | ref | (.) | ref | (.) |
| Booster-vaccinated | 0.72 | (0.56-0.92) | 0.71 | (0.55-0.93) | 0.62 | (0.47-0.83) |
| **Primary case age** |  |  |  |  |  |  |
| 0-10 | 1.28 | (1.10-1.50) | 1.44 | (1.22-1.69) | 1.53 | (1.29-1.82) |
| 10-20 | 1.04 | (0.90-1.21) | 1.14 | (0.98-1.33) | 1.19 | (1.01-1.41) |
| 20-30 | ref | (.) | ref | (.) | ref | (.) |
| 30-40 | 1.92 | (1.65-2.25) | 1.93 | (1.64-2.28) | 2.02 | (1.69-2.42) |
| 40-50 | 2.31 | (1.98-2.70) | 2.45 | (2.08-2.88) | 2.45 | (2.05-2.93) |
| 50-60 | 2.45 | (2.09-2.87) | 2.64 | (2.24-3.11) | 2.72 | (2.26-3.26) |
| 60-70 | 3.16 | (2.54-3.94) | 3.22 | (2.56-4.06) | 3.13 | (2.43-4.02) |
| 70+ | 3.99 | (2.76-5.75) | 4.11 | (2.82-6.00) | 3.64 | (2.40-5.52) |
| **Potential secondary case age** |  |  |  |  |  |  |
| 0-10 | 0.82 | (0.71-0.95) | 0.89 | (0.76-1.03) | 0.94 | (0.79-1.11) |
| 10-20 | 0.70 | (0.61-0.80) | 0.73 | (0.63-0.84) | 0.75 | (0.64-0.88) |
| 20-30 | ref | (.) | ref | (.) | ref | (.) |
| 30-40 | 1.64 | (1.44-1.88) | 1.70 | (1.47-1.96) | 1.77 | (1.51-2.08) |
| 40-50 | 1.60 | (1.40-1.82) | 1.67 | (1.45-1.92) | 1.69 | (1.45-1.98) |
| 50-60 | 1.48 | (1.29-1.71) | 1.48 | (1.28-1.72) | 1.55 | (1.32-1.84) |
| 60-70 | 1.53 | (1.25-1.87) | 1.59 | (1.29-1.95) | 1.58 | (1.25-1.99) |
| 70+ | 1.01 | (0.75-1.37) | 1.03 | (0.75-1.41) | 1.08 | (0.76-1.54) |
| **Household size** |  |  |  |  |  |  |
| 2 | 2.26 | (1.85-2.75) | 2.27 | (1.85-2.79) | 2.28 | (1.82-2.85) |
| 3 | 1.53 | (1.26-1.86) | 1.52 | (1.24-1.86) | 1.57 | (1.26-1.96) |
| 4 | 1.51 | (1.25-1.82) | 1.52 | (1.25-1.84) | 1.64 | (1.33-2.03) |
| 5 | 1.40 | (1.15-1.70) | 1.43 | (1.17-1.76) | 1.54 | (1.23-1.93) |
| 6 | ref | (.) | ref | (.) | ref | (.) |
| **Primary case sex** |  |  |  |  |  |  |
| Male | ref | (.) | ref | (.) | ref | (.) |
| Female | 0.99 | (0.92-1.07) | 1.00 | (0.93-1.08) | 0.97 | (0.89-1.06) |
| **Potential secondary case sex** |  |  |  |  |  |  |
| Male | ref | (.) | ref | (.) | ref | (.) |
| Female | 1.14 | (1.07-1.20) | 1.14 | (1.08-1.21) | 1.14 | (1.06-1.21) |
| Number of observations | 27,874 |  | 27,011 |  | 25,611 |  |
| Number of households | 11,937 |  | 11,709 |  | 11,338 |  |

Notes: This table shows the sensitivity for the choice of inclusion of positive secondary cases. In Table 9 we show the estimates, while restricting the inclusion criteria for positive secondary cases. Column I shows the odds ratio estimates, including positive secondary cases found on day 1-7, i.e., our main specification, also presented in Table 2 and 3. Column II includes positive secondary cases found on day 2-7. Column III includes positive secondary cases found on day 3-7. 95%-Confidence interval in parentheses. Standard errors are clustered on the household level.

Column I of Table 8 shows the odds ratio estimates of our main specification, i.e., the same as used in Table 2 and 3. Column II shows the odds ratio estimates, when excluding households with a primary case <10 years. Column III shows the odds ratio estimates, when only including 2-person households. Column IV shows the odds ratio estimates, while adjusting for Ct value of the primary case.

In Table 9 we show the estimates, while restricting the inclusion criteria for positive secondary cases. Column I shows the odds ratio estimates, including positive secondary cases found on day 1-7, i.e, our main specification, also presented in Table 2 and 3. Column II includes positive secondary cases found on day 2-7. Column III includes positive secondary cases found on day 3-7.

